# Developing a Population Health Management Dashboard for Belgium: a focus group study with mock-up design

**DOI:** 10.1101/2025.04.25.25326410

**Authors:** Marie Van de Putte, Laura Christiaens, Laura Goetschalckx, Stefan Morreel, Joeri R. Verbiest, Mare Claeys, Josefien Van Olmen, Bert Vaes, Liesbet M. Peeters

**Affiliations:** Department of Public Health and Primary Care, KU Leuven, Kapucijnenvoer 7 bus 7001 blok h, 3000 Leuven, Belgium; Zorgzaam Leuven, Maria Theresiastraat 63/A, 3000 Leuven, Belgium; Department of Primary and Interdisciplinary Care Antwerp Research Group, University of Antwerp, Antwerp, Belgium; Biomedical Research Institute (BIOMED), Hasselt University, Agoralaan Building C, Diepenbeek, Belgium; Data Science Institute (DSI), Hasselt University, Agoralaan Building D, Diepenbeek, Belgium; Centre for Research in Health System Performance (CRiHSP), Yong Loo Lin School of Medicine, National University of Singapore. Department of Family Medicine (DFM) National University Health System

**Author notes:** Correspondence to: Liesbet Peeters, Address: Agoralaan Building C, 3590 Diepenbeek, Belgium.

**Keywords:** Population Health Management, Dashboard, Dashboard requirements, User need analysis, Focus groups, Health data integration, Health indicators, Risk stratification, Intervention selection

## Abstract

**Background:** The necessary data for delivering targeted population health management (PHM) interventions in Belgium is insufficient and scattered due to limited, non-interoperable data sources. Insights on data availability, user needs, and analysis capabilities should be obtained to develop a Belgian PHM dashboard. We aimed to identify visual, content, and access requirements for a PHM dashboard, and visualize user needs through a mock-up dashboard to enhance quintuple aim.

**Methods:** Five focus groups were conducted in three Belgian locoregional health networks (collaborations of primary care zones), comprising 35 potential future dashboard users with various backgrounds. Insights were gathered using open-ended questions based on frameworks of successful international examples. These insights were incorporated into a mock-up dashboard. Subsequently, the mock-up dashboard was discussed in a second round of focus groups. Data from the focus groups were analysed using the Qualitative Analysis Guide of Leuven (QUAGOL) method.

**Results:** Differentiated user requirements can be classified into: (1) ’General Exploration’ allowing for assessing the health and socioeconomic status and risk factors in a region using PHM indicators, (2) ‘Risk Stratification and Selection of Interventions’ enabling matching of resources and interventions to at-risk subpopulations, (3) ‘Research Community’ providing advanced tools for data exploration and collaboration with the research community. The mock-up dashboard meets these requirements according to respondents, by supporting population health managers, healthcare professionals and policymakers with easy access to insights and expert population health managers, such as researchers and data analysts, with options for advanced data analysis.

**Conclusions:** Our study identified ‘exploration’, ‘risk stratification’, ‘intervention selection’ and ‘advanced data analysis’ as key components of an interoperable and customizable dashboard, contributing to a more efficient and equitable healthcare system. The developed PHM mock-up dashboard encapsulates these features, aiming to achieve the quintuple aim in Belgium.

## Background

’Integrated care’ is a multidimensional and continuously evolving concept representing a health system approach that addresses the multidimensional needs of the entire population (1). The effective utilization of data, encompassing health-related data, social determinants of health, and environmental factors, is crucial for achieving integrated care and performing population health management (PHM) (2). PHM is described as a people-centred, data-driven and proactive approach to manage the health and well-being of a defined population, considering the differences within that population and their social determinants of health (3). Beyond enhancing the quintuple aim (5AIM) outcomes, PHM seeks to more precisely delineate the health system’s priority actions (4–6). Examples of PHM systems include ‘Affordable Care Organizations’ in the United States of America, *‘Gesundes Kinzigtal*’ in Germany, and the ‘*Extramuraal Leidens universiteir Medisch Centrum Academisch Netwerk*’ in the Netherlands (7–9).

To integrate the required data in a user-friendly manner, dashboards are crucial for transforming complex data into actionable insights via interactive visual interfaces, enhancing the utilization of data in care delivery (10–13). By summarizing performance indicators, displaying predictive modelling results, benchmarking against peers, and flagging measures against target values, dashboards enable continuous health monitoring, disease outbreak prediction and management, quality of care improvement, and effective resource use (10,11,13–16). In developing dashboards, it is essential to select data following the FAIR (Findable, Accessible, Interoperable, and Reusable) principles, ensure the interoperability of consulted data sources, and define analysis methods and interpretation standards to visualize complex and heterogeneous data in a user-oriented manner (17–21). To maximize their effectiveness, these dashboards and their outputs should be available via open-source software and customizable to accommodate user needs (22). Examples illustrating how different elements can be integrated to meet all requirements for a PHM dashboard include the U.S. City Health Dashboard, the Nottingham PHM Blueprint and the European Health Data Evidence Network (EHDEN) Portal (23–25). Development should also consider the various barriers associated with implementing such an integrated dashboard, including stakeholders’ benchmarking fear, insufficient engagement of citizens and stakeholders in PHM scientific reports, IT burdens and difficulties in using the tools (26–28).

Despite the existence of disease-specific and multipurpose dashboards such as the dashboards of the InterMutualistic Agency and Provincie in Cijfers in Belgium, the country lacks a comprehensive dashboard that captures the various dimensions required for PHM and cannot integrate diverse data types from multiple sources due to the fragmented and non-interoperable data landscape (1,29–31). This makes it impossible to collect and analyse PHM data holistically. Furthermore, the development of a national integrated PHM dashboard is impeded by Belgium’s complex state structure, comprising six governments and nine health ministers from both federal and federated entities (32,33). In 2023, a Memorandum of Understanding was signed between Belgian federal government and federated entities, greenlighting the launch of a new Inter-federal Plan for Integrated Care in Belgium to develop a population management strategy (34).

To assist policymakers in creating a PHM dashboard in Belgium, the Pilot Research On a Scalable Population Health Management Connected Dashboard (PROSPeCD) initiative was launched. Supported by the Data4PHM consortium, it aimed to identify content and access requirements for a PHM dashboard based on user needs, explore meso-level designs, and address implementation facilitators and barriers (35). The goal was to create a Belgian PHM mock-up dashboard as a scalable, interoperable example to enhance 5AIM outcomes and facilitate integrated care and PHM use at the neighbourhood (micro), locoregional (meso), and national or federated entities (macro) levels.

## Methods

To comprehensively understand the visual and content-related needs of potential PHM dashboard users, as well as to identify PHM implementation barriers and facilitators, a qualitative design using focus groups was selected. Focus groups foster dynamic dialogues and promote an in-depth understanding of the needs of potential users (36). Concurrently, a mock-up dashboard – a visual representation of the intended dashboard – was developed using an iterative, user-centred approach to test user requirements and technical feasibility with end users.

### Study design and population

Five focus groups were conducted in Belgium (Flanders) across three locoregional health networks: *Eerstelijnszones Antwerpen* (three primary care zones (PCZs), a local network of healthcare providers and social workers that integrates care and well-being services to meet the needs of the community), *Zorgzaam Leuven* (six PCZs), and *Centrum Eerste Lijn Limburg* (CELL) (eight PCZs). Each locoregional health network provided a list of potential PHM dashboard users operating within their respective PCZs. To ensure the requisite number and enhance the diversity of participants, researchers contacted additional potential participants. They were all contacted via email to ask for their participation. Participants were included if they met the inclusion criteria while considering the desired diversity of the panel in terms of age, gender, years of experience, and profession (see Supplemental material 1). Participants were paid €100 per focus group.

### Data collection

Five focus groups, each with a maximum duration of 120 minutes, were conducted live or via MS Teams between May and September 2024. Each focus group was moderated by a PHM expert, supported by another expert in PHM as observer. These PHM experts were members of the Data4PHM consortium: authors MVDP (pharmacist and post-doctoral researcher), LG (general practitioner (GP) in training and student researcher), MC (pharmacist and population health manager) and SM (GP and post-doctoral researcher). Most of the participants knew the experts and were aware of their enthusiasm for local population management. For clarity, the researchers and their objectives were introduced at the beginning of the focus groups.

Focus groups were organized in two rounds. Round one, organized in locoregional health networks *Eerstelijnszones Antwerpen* and *Zorgzaam Leuven,* collected needs of potential PHM users regarding dashboard effectiveness (e.g. usability, accessibility, interoperability, practical application, and feedback strategies), and end-user involvement. Round two, organized in all locoregional health networks, captured feedback and perceptions of PHM dashboard users based on an integrated, user-centred PHM mock-up dashboard.

The experts utilized topic guides to structure the two rounds of focus groups, ensuring a consistent approach across the different locoregional health networks (see Supplemental material 2). The guides included open-ended questions based on theoretical frameworks, such as Steenkamer’s Collaborative Adaptive Health Network (CAHN) framework (37). Based on the first round’s preliminary results, the second round’s topic guide was modified. All focus groups were audio or video recorded and transcribed verbatim with the participants’ consent. Due to time and resource limitations, these transcripts were not returned to the participants.

### Analysis

Data analysis was conducted using the Qualitative Analysis Guide of Leuven (QUAGOL), a practice-based guide that supports and facilitates the analysis of qualitative interview data (38). This method comprises two phases, each consisting of five stages: the preparation of the coding process (using paper and pencil), and the actual coding process (supported by the qualitative software program QSR NVivo 14).

In the initial phase, raw data were reviewed to develop an empirically based conceptual framework, which provided relevant concepts for answering the research question and served as the foundation for coding and analysis. Subsequently, these concepts were critically evaluated, integrated and reformulated in the second phase to define a conceptual framework. The method’s iterative approach, involving continuous movement between different stages and within-case and across-case analyses, facilitated a deeper understanding of the results and verification of hypotheses with newly collected data.

The reliability of the analysis was ensured by maintaining narrative review reports, conceptual interview schemes, code trees, and analysis schemes. All focus groups were double-coded by two researchers (LC, LG), and analysed by the research team (LC, LG, SM, MC, MVDP).

### Mock-up dashboard development

The first version of the mock-up dashboard was developed based on three key sources: 1. desired features identified during the first round of focus groups, 2. insights from current literature on PHM, and 3. ad-hoc web searches. Expert support was obtained from specialists in data science, PHM, and leaders from exemplary initiatives. These preliminary concepts were presented to potential PHM dashboard users during the second round of focus groups, using example visualizations with fabricated data to illustrate potential functionalities and user interfaces. The outcomes from the second round of focus groups facilitated the refinement of the dashboard’s features, underscoring the iterative co-creation process fundamental to its development.

### Ethical consideration

The ethical committee of the University of Antwerp approved the study design and waived written consent (Project ID 6554). All participants were sent an Informed Consent Form detailing the research in advance. At the commencement of the focus group, verbal consent was obtained and recorded.

## Results

In total, 35 participants (19 women and 16 men) participated in the five focus groups. The age of the participants ranged from 20 to 64 years and their experience from 0 to 34 years (***Error! Reference source not found.***).

Focus groups on PHM dashboard requirements identified three interconnected themes with subthemes: data (availability, quality, interoperability), users (training in dashboard use, dashboard accessibility, role), and analysis (population health analysis, intervention selection, stratification) (Figure 1) (see Supplemental material 3). Periodic evaluation of each theme is necessary. The mock-up dashboard can be consulted electronically (<u>PROSPeCD_mock-up_dashboard</u>).

**Table 1.** Participants’ personal characteristics (N = 35)

| Gender | Male |  |  | Female |  |  |  |  |  |
| --- | --- | --- | --- | --- | --- | --- | --- | --- | --- |
|  | 16 |  |  | 19 |  |  |  |  |  |
| Age category | 20-24 | 25-29 | 30-34 | 35-39 | 40-44 | 45-49 | 50-54 | 55-59 | 60-64 |
|  | 1 | 3 | 3 | 6 | 8 | 4 | 3 | 6 | 1 |
| Years of experience | 0-4 | 5-9 | 10-14 | 15-19 | 20-24 | 25-29 | 30-34 |  |  |
|  | 3 | 5 | 4 | 8 | 8 | 2 | 5 |  |  |
| Profile of potential dashboard users | Healthcare professional |  | Healthcare manager |  |  | Policy maker |  | Data expert |  |
|  | General practitioner | 7 <sup>(*2)</sup> | Community health service employee |  | 2 <sup>(*1)</sup> | City official | 1 | Data expert | 3 <sup>(*1)</sup> |
|  | Nurse | 1 <sup>(*1)</sup> | Implementation manager of integrated care |  | 4 | Implementation manager of integrated care | 4 <sup>(*2)</sup> |  |  |
|  | Community pharmacist | 6 <sup>(*3)</sup> | Hospital process manager |  | 2 | Primary care zone council | 3 |  |  |
|  |  |  | Population health manager |  | 2 <sup>(*1)</sup> |  |  |  |  |
|  |  |  | Representatives of primary care profession |  | 7<sup>(*2)</sup> |  |  |  |  |

*\*Number of participants with dual profiles*

**Figure 1.**
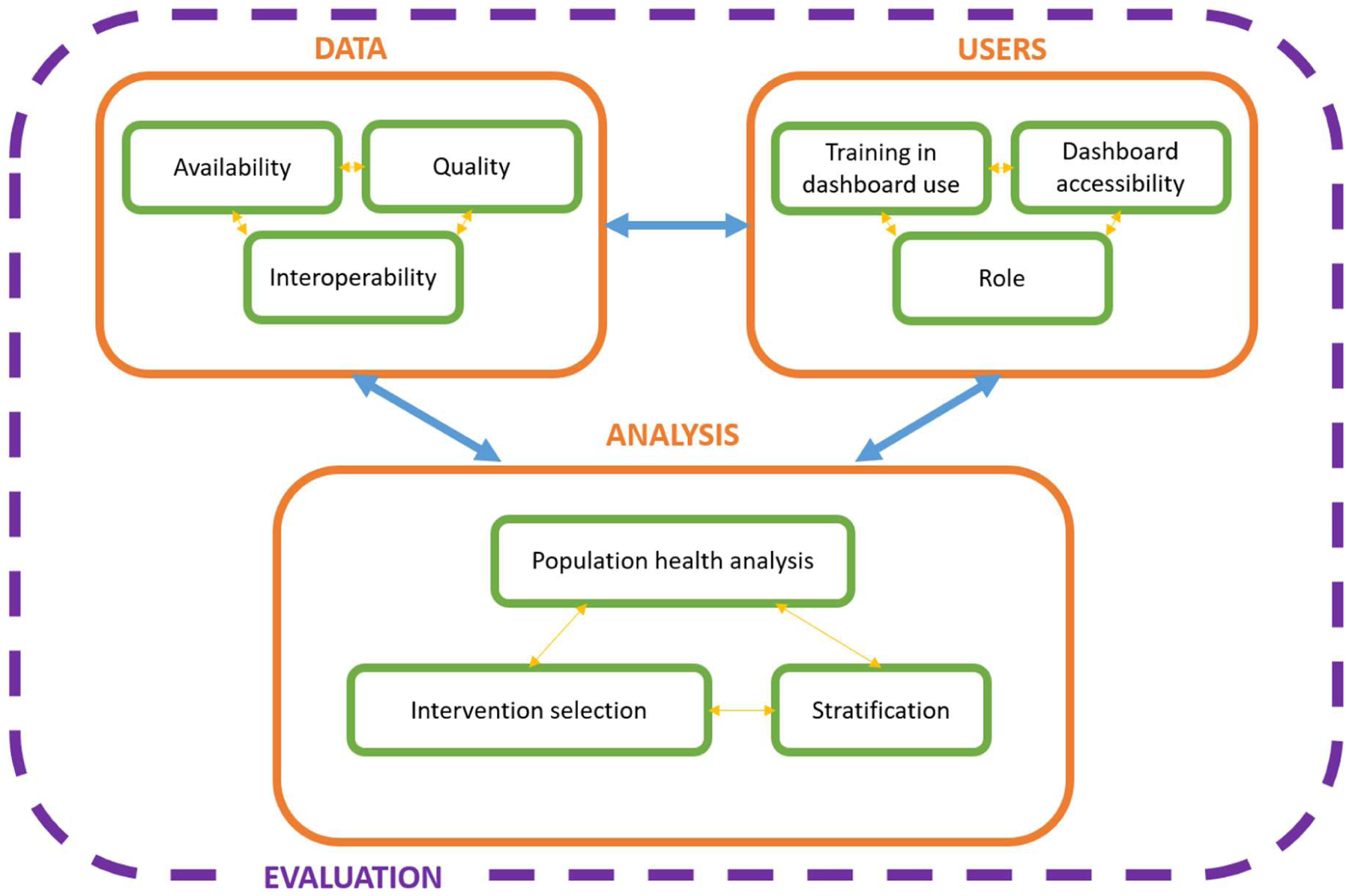
Interconnected (sub)themes derived from focus groups. Requirements for a population health management dashboard are categorized into three interconnected themes: data, users, and analysis.

### Availability, Quality and Interoperability of Data

Participants emphasized the need for the dashboard to encompass diverse data by defining clusters of metrics for general exploration. Recommended clusters included health and well-being, socioeconomic, health behaviour, environmental, and clinical care outcomes. Metrics should be defined in both relative percentages and absolute numbers to contextualize data and provide accurate statistics for policymaking. Each outcome required a clear definition, data source references, and raw data availability. Governance is crucial for selecting new themes and associated data.

> *“If you can’t give clear statistics to policymakers, they will not take your policy advice seriously.”*
>
> *(Community health service employee)*

Data should be available at various levels, including neighbourhoods, PCZs, and (local) government levels. Integrating macro and meso levels with the micro level allowed data filtering by the required level. Participants preferred data corresponding to their practice’s operating area for tailoring local health interventions and adjusting policies. However, they indicated that limited coding options in the EHR and other data sources restrict the structural inclusion of data in the dashboard.

Up-to-date and densely collected data was crucial, as unreliable or outdated data undermined the dashboard’s credibility and sustainable use. Enhancing registration quality could be supported by involving data providers in dashboard development and requiring data input in a format immediately usable for dashboards.

> *“The dashboard should be designed to evolve over time as new insights emerge. Additionally, it is crucial to keep the data as up-to-date as possible.”*
>
> *(Primary care zone staff / population health manager)*

Barriers to data sharing included concerns regarding the preparedness and willingness of data partners to share information. Additionally, data at the lowest possible geographical level is desired, as this enables the execution of small cell risk analyses, which are crucial for implementing adequately targeted local healthcare interventions. However, this necessitates a sufficiently dense data collection at this low geographical level; otherwise, it may comprise the privacy of data providers, with the risk of individual practices being identifiable. Further barriers included GDPR regulations, population reluctance to share data, and withdrawal of consent for data sharing.

A federated data network approach is recommended, which combines information from hospitals, GPs, pharmacists, paramedics, and other healthcare providers in a collaborative manner. However, it is crucial to establish a consistent dashboard architecture with stable data definitions over time across different regions to ensure interoperability. This may involve defining a national architecture for the dashboard, along with individual interfaces for each federated entity, tailored to their specific focus.

### Dashboard users

Participants emphasized the necessity for the dashboard to be adaptable to various users, rather than being restricted to specific job titles. Numerous individuals, encompassing staff from PCZs, local governments, hospital process managers, and integration managers, were already undertaking PHM responsibilities, yet were hindered by absence of suitable tools.

> *“Rather than being a specific job, population management is a role that various individuals can fulfil. Today, many individuals are already embracing this responsibility in their current role.”*
>
> *(Population health manager)*

At the practice level (e.g. primary care practice, community pharmacists), profiles with a PHM function were expected to analyse data, visualize results, and determine focus areas. Population health managers at the neighbourhood or meso level is expected to manage the dashboard, send notifications when issues arose, and collaborate with healthcare and welfare workers to explain data findings, gather neighbourhood team experiences, and evaluate potential causes. Clinical support managers should be able to create strategic care plans using dashboard data, while PCZs collaborate with health professionals and local governments for interventions. Data analysts need to conduct in-depth analysis to support dashboard users, perform impact analyses, and interpret data. Policymakers are responsible to monitor domain-specific data to guide policy decisions, prioritize interventions based on parameter alarms, and manage the dashboard at the governmental level. However, data providers feared benchmarking, as the government could use the data to control or punish them. Cooperation among stakeholders was stipulated as being essential to gain in-depth knowledge about themes and the population, and to map the context behind the data.

Participants indicated that the dashboard should follow a dual approach to address diverse user needs, consisting of a publicly available space (PHM dashboard) and a closed, login- protected, environment for researchers and data experts (PHM platform). The PHM dashboard would be accessible to anyone interested in the data, offering general exploration and risk stratification features. The PHM platform would require sign-in (preferably single sign-on from medical software for healthcare professionals) and extensive expertise in data sciences to mitigate information overload and prevent misinterpretation. This setup would enable comprehensive analysis to generate new population health insights, collaborative research, data sharing, and pilot testing of new features before integrating them into the publicly accessible PHM dashboard.

Geographic levels (e.g., PCZ, neighbourhood) and data types (e.g., person characteristics and health data, socioeconomic data, environmental factor) should be adjustable according to user roles. Training through e-learning modules, incorporating examples and exercises tailored to user profiles, was essential to ensure accurate data interpretation and prevent miscommunication by users with insufficient knowledge.

> *“Describe high needs well, so that a layman in dashboard use does not think that he has to focus on it 100%, while we actually want to keep the strata and focus on prevention. It is not like ‘oh it is red, I only have to focus on that.’”*
>
> *(Primary care zone staff / community pharmacist)*

### Analyses through the dashboard

#### General exploration

The dashboard facilitated the delineation of the population both in general terms and according to specific themes. It identified highlights based on deviating parameters and support users in confirming or refuting their findings.

> *“On the one hand, the dashboard must be proactive, the data should give you important information that you would not look for yourself; the ‘blind spot’. On the other hand, it must connect with what is felt in the neighbourhood, so that it can be confirmed by data.”*
>
> *(Population health manager)*

Highlights were generated when deviating parameters were noticed, based on predefined criteria. These included key indicators and comparisons on health, social and economic parameters for the chosen region, the most affected regions for chosen themes, and identified blind spots. Users could customize additional highlights based on their goals, target groups, and themes.

> *“It is important to be able to ‘turn it on’. We have developed a number of systems like this before …, but the pitfall is that there are too many pop-ups and too many systems going off. So it does have to be dosed, …, so you can see what we need to focus on.”*
>
> *(General practitioner)*

Users analysed themes in relation to regions or target groups (Figure 2a/b), gaining insights into profiles behind metrics, identifying blind spots, and integrating findings with contextual knowledge. They would have the ability to add filters to maps, benchmark against other

**Figure 2.**
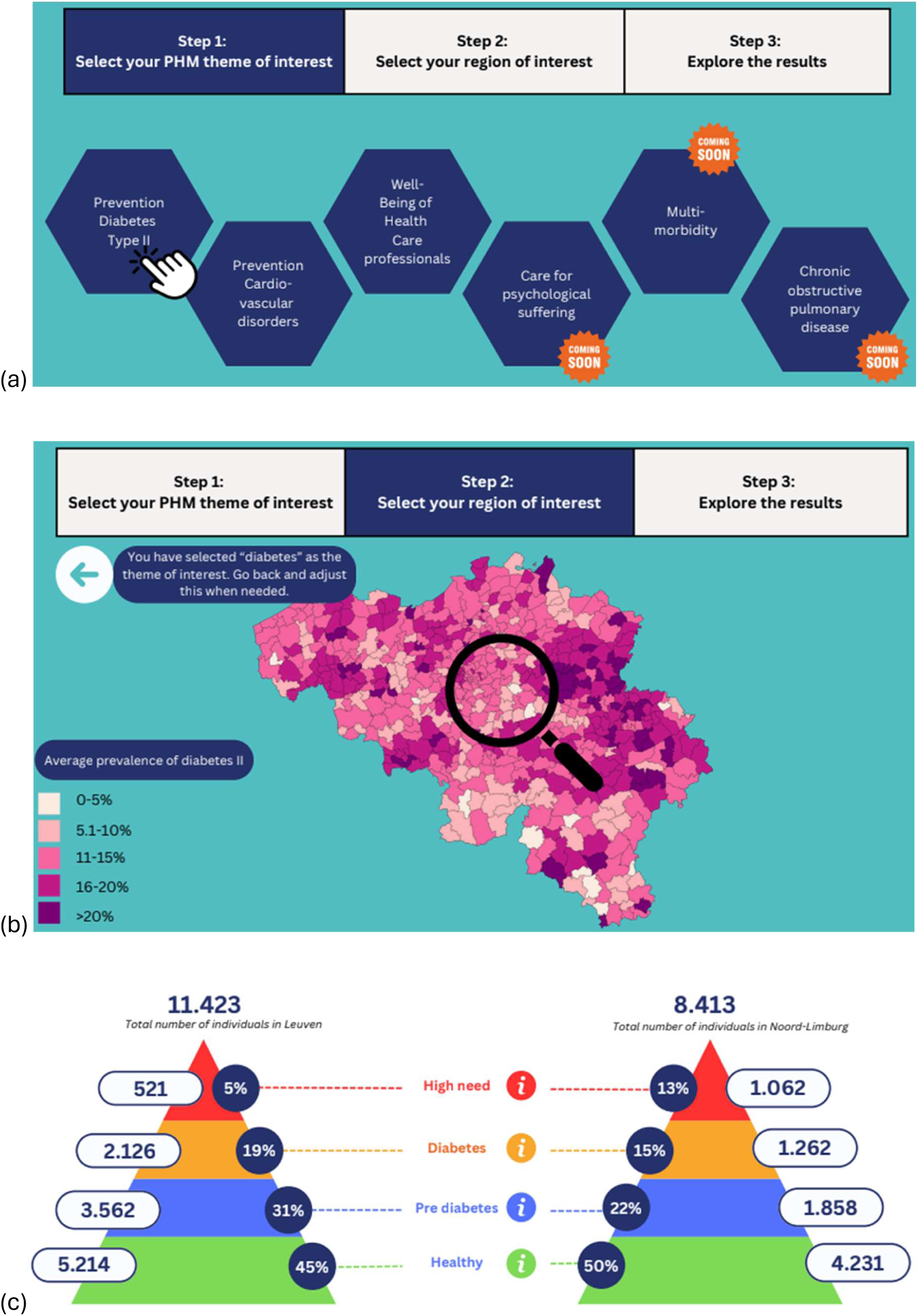
Guided Population Health Analysis Using a Mock-up Dashboard. (a) Population health management (PHM) dashboard users can conduct theme-specific explorations of population health; (b) Based on the selected PHM theme, users can choose a region of interest, providing both visual and quantitative representation of the theme’s prevalence; (c) A theme- and region-specific period represents the distribution of individuals across strata, which can be benchmarked against comparable regions using adjacent pyramids representing the selected and comparator regions; (d) A dynamic graph visualizes historical trends of the strata, enabling the observation of the health issue’s evolution, identification of emerging patterns, and detection of declines in prevalence rates. (e) Information buttons are included to allow users to access additional details on time trends and intervention selection.

(international) cities or neighbourhoods with similar demographic (Figure 2c) and/or social characteristics and analyse trends over time (Figure 2d) by comparing parameters across different levels or regions over time. Impact evaluations were based on actions and simulations, using metrics like quality-adjusted life years, adherence rates, and increases in evidence-based activities. Users would also need to be able to analyse care programs and determine optimal budget spending.

Facilitators for data exploration included an ‘info’-button (Figure 2e) for correct data interpretation (e.g. time frame of data collection), artificial intelligence (AI)-supported insights and visualizations, and features like graph legends, sliders, search functions, and saveable modular layouts tailored to user needs. The dashboard was designed to be user-friendly, with an intuitive interface with low cognitive load requiring no specialized education in data science.

> *“Since most users won’t have a data science background, it’s essential that the interface is very intuitive. Additionally, only ‘validated insights’ should be shared with such a wide audience.”*
>
> *(Population health manager)*

Barriers to data analysis included missing contextual data, leading to incorrect impact evaluations, and political cherry-picking. Highlights alone were insufficient without additional support and solutions. Obtaining supplementary information often required consulting other websites, creating a barrier to efficient analysis. Data was not immediately available in a suitable format, necessitating multiple steps to access the required information. The risk of assessment evolving into a funding instrument could trigger gaming, undermining data integrity.

#### Stratification

Users could stratify the population by theme into strata based on their personal characteristics, health and/or socioeconomic status, or environmental factors. The number of strata was theme specific. A pyramid representation, similar to the Kaiser Permanente Triangle, was considered feasible and intuitive, aiding in detecting switches of strata, investigating causes and context, and predicting population evolution within the pyramid (39).

> *“How quickly do people move from one stratum to another, and what are triggers for this?”*
>
> *(Data-expert)*

AI integration into the dashboard was proposed for tasks like visualization and stratification. However, this integration was only recommended if scientifically defined algorithms, established through the Delphi method, were used and complemented by human review to validate classifications, considering contextual information.

#### Intervention selection

Metrics deviating from predefined cutoff values should be flagged through dashboard alerts. This insight could subsequently serve as a foundation for selecting and prioritizing interventions. A multidisciplinary expert panel, including scientists, local policymakers, and dashboard users, is best placed to suggest interventions. Those interventions, targeting (sub)populations within the stratification triangle, were proposed to address challenges identified through data and blind spots, considering budget, system burden, number needed to treat, and return on investment. Suggestions included already implemented interventions with context and impact information for evaluating relevance in new contexts. These suggestions encompassed a combination of both national and local initiatives linked to prevention workers for support. Defined actions needed to be feasible and evidence-based to avoid redundancy.

Facilitators to intervention selection included providing expert support to low-scoring care centres instead of financial punishment. The government was advised to allocate a 2-3% budget margin for targeted efforts based on dashboard data, empowering users to analyse and act on the data. The impact of proposed interventions was indicated using past similar situations. Barriers included potential conflicts with local policies and the risk of misidentifying individuals for intervention support.

### Dashboard evaluation

The dashboard evaluation should be periodically reviewed by both current and past dashboard users, as well as developers, to ensure its operability. This should include feedback provision on data availability, the (un)availability of analytical capabilities, and mapping out the reasons for both the use and discontinuation of the dashboard.

## Discussion

This initial framework on required user needs (general exploration, risk stratification and intervention selection, and research community), and a mock-up dashboard, serves as a foundational base for future PHM dashboard development and a testbed for refining functionalities, ensuring alignment with user needs and technical standards.

### User needs

General exploration allows users to broadly explore data across regions, offering a comprehensive overview of health, social, and environmental outcomes to support a holistic understanding of region’s population health and wellbeing (2,40). The desired indicators, analytical functionalities of the metrics (e.g. assessment of alignment or divergence across regions), supporting tools such as tips and cautions on using the data, and visual elements like color-coded evolution of parameters, are consistent with the format of the U.S. City Health Dashboard (23). In Belgium, resources like ’*Provincies in Cijfers*’ provide detailed regional data, reports, regional comparisons, and analysis tools, aiding local governments in making informed, data-driven decisions (30). However, data aggregation to support this demographic overview is challenging due to complex geographical data structures and non-standardized regional classifications. Therefore, data harmonization is desirable as it consolidates data from various sources while preserving context and ensuring a consistent and standardized meaning. This facilitates interoperability allowing data to be easily exchanged, combined, and analysed seamlessly. Adopting the ‘Observational Medical Outcomes Partnership Common Data Model (OMOP-CDM)’ helps overcome those challenges by standardizing and simplifying data integration from diverse sources, thereby supporting a scalable infrastructure (41). In Belgium, initiatives like the Belgian eHealth platform promote electronic data exchange and support international standards like Fast Healthcare Interoperability Resources (FHIR), which facilitate interoperability but are unsuitable for large-scale data analytics (42).

Data-driven insights allow population stratification by health or social risk, guiding interventions aligned with Belgian standards and optimizing resource allocation. The ‘supporting selection of interventions’ feature lists potentially suitable interventions for specific (sub)populations based on international guidelines and expert knowledge, preventing redundant efforts. Post-implementation, impact evaluations can be conducted. Although risk stratification and impactibility modelling may be biased towards vulnerable groups, they enhance access and equity (43). However, stigma and potential misclassification pose significant ramifications (44).

The Research Community requires advanced capabilities for deep data exploration and collaborative research. The dashboard provides a detailed, downloadable metadata catalogue to ensure the findability of PHM-relevant data sources, allowing users to tailor their analysis or apply customized methodologies that are not (yet) available within the dashboard. Introducing this feature requires careful consideration to address privacy concerns and maintain data integrity and security. Data custodians’ concerns about public data sharing must be acknowledged, ensuring all original data sources permit sharing under clearly defined terms of use. Following the example of the U.S. City Health Dashboard, these terms of use should include clear disclaimers and cautions to inform users of any usage restrictions before data download (23).

### Federated data

Depending on information needs, analyses can be federated or centralized. Federated analysis preserves privacy and data control by analysing data locally, though it limits patient-level data linking. Emerging technologies like secure multi-party computation and federated learning may address these limitations. Centralized analysis offers deeper insights by combining data from multiple sources but poses higher privacy and security risks and reduces data provider control. Thus, the dashboard should prioritize a federated approach, as used in the EHDEN portal, to balance insights with privacy, using centralized analysis only when its benefits outweigh the risks (25). However, designing a federated dashboard is challenging due to diverse data sources and formats, resulting in low interoperability and reusability (1,29).

### Dashboard reliability

By alerting users to potential biases or data limitations, the dashboard ensures data integrity and responsible use. A help desk assists with complex data navigation, reducing errors, and enhancing user satisfaction. Next, users fear benchmarking when government-managed dashboards may lead to penalties for poor performance (26). Therefore, the PHM dashboard should be positioned as a tool for health improvement and motivation, not for assessing healthcare providers. The user policy should explicitly state that dashboard results will never guide individual financing or evaluate healthcare professionals’ performance. In the context of PHM, this issue can be addressed by developing indicators and benchmarks based on consensus among care providers at various levels (26).

### Quality improvement and expansion of the dashboard

Iterative feedback loops are essential for developing the PHM dashboard, ensuring new functionalities align with real-world applicability, user requirements, and technical feasibility (26,27). Your insights and feedback on our dashboard, accessible via <u>PROSPeCD_mock-up_dashboard</u>, are valuable for the continued refinement of the PHM dashboard. The dashboard should evolve by incorporating new themes and indicators selected by multidisciplinary PHM expert teams, aligning with user expectations and public health priorities. Prioritizing programs that align with the Interfederal Plan for Integrated Care—Perinatal Care, Vulnerable Populations, and Childhood and Youth Obesity—would be beneficial (34). However, governance must be established to define criteria for selecting new themes.

A controlled demo environment with restricted access should be used within the PHM Platform to ensure accurate and interpretable data visualization, allowing advanced PHM and data science users to rigorously test and refine new features and technical requirements, ensuring clarity and trustworthiness.

The feasibility of the dashboard’s successful implementation in daily practice is promising due to several factors. Belgium’s extensive data resources, including InterMutualistisch Agenschap – Agence InterMutualiste for healthcare utilization, Statbel for socio-economic indicators, Zorgatlas, Intego and Farmaflux, provide a robust foundation for a unified PHM effort. Focused program priorities within the Interfederal Plan for Integrated Care offer clear guidance for theme and indicator selection. Existing expertise and collaboration among consortia and experts, supported by a shared data infrastructure, can reduce costs and enhance efficiency. The supportive technological landscape, including the European Health Data Space and Belgium’s Health Data Agency, facilitates scalable technology solutions. Adopting a common data model like OMOP CDM ensures interoperability and data integration while maintaining privacy.

### Strengths and limitations

Participation from three locoregional health networks provided valuable insights into varied user needs for the PHM dashboard. However, the recruitment process may have introduced bias by selecting enthusiastic users, potentially skewing results towards more complex data analysis functionalities than the average potential dashboard user possesses in terms of handling and data interpretation capabilities. Future research should establish locoregional health networks in other Belgian regions for broader geographic representation.

Focus groups with open-ended questions facilitated detailed responses and observations of user interactions, offering insights into collaborative discussions about the PHM dashboard. Nevertheless, this method does not allow for the quantification of the relative importance of different requirements.

## Conclusions

By mapping the needs of potential PHM dashboard users regarding general exploration, risk stratification and intervention selection, and a research community, a comprehensive framework was developed for a scalable, secure, and interoperable PHM dashboard. Customizing the dashboard will facilitate the creation of interventions tailored to the population’s needs while aligning with regional healthcare priorities. Consequently, the dashboard will support 5AIM health outcomes and facilitate integrated care at neighbourhood (micro), locoregional (meso), and national (macro) levels, thereby advancing PHM and integrated care in Belgium.

## Data Availability

The raw data will not be shared to protect the identity of participants. The dataset supporting the conclusions of this article is included within Supplemental material 3. Requests for de-identified raw data will be considered by the authors. A more detailed overview of the mock-up dashboard and the opportunity to provide feedback is available via the link.

https://zenodo.org/records/15165445

## Declarations

### Ethical approval and consent to participate

Ethical approval for the study was obtained from the Ethics Committee of the University of Antwerp (Project ID 6554). Prior to the study, all participants received an Informed Consent Form outlining the research details. At the start of the focus groups, verbal consent was obtained and recorded. These consent recordings will be securely stored for 20 years. Participants were assured that no potentially identifying details would be reported, all data would be kept confidential, and they could withdraw at any time or decline to answer any question without any negative consequences.

### Availability of data and materials

The raw data will not be shared to protect the identity of participants. The dataset supporting the conclusions of this article is included within Supplemental material 3. Requests for de-identified raw data will be considered by the authors. A more detailed overview of the mock-up dashboard and the opportunity to provide feedback is available via the following link: <u>PROSPeCD_mock-up_dashboard</u>

## Funding

This study is funded by the EU’s Recovery and Resilience funds allocated to Belgium for digital health transformation and developed within the EU Technical Support Instrument (TSI) ‘EU Resources Hub for Sustainable Investing in Health’, managed by the Commission’s Directorate General for Structural Reform Support providing demand-driven, customized technical expertise for member states to design, plan and implement reforms.

### Author’s contributions

All authors read and approved the final manuscript.

## Acknowledgements

The authors would like to thank all participants of the focus groups for their time and contributions. For the locoregional health network in Limburg, we extend our sincere gratitude to Anouk Geenen and Anouk Tuinstra for their observations and support during the focus groups. We would also like to thank the steering committee members, including Annemarie Jacobi, Ann Marie Borg and Andrew Amato.

