## Supplemental material for "Developing a Population Health Management Dashboard for Belgium: a focus group study with mock-up design"

### Supplemental material 1 Inclusion criteria focus group participants

#### 1. ≥ 18 years old

#### 2. Participants must be potential user of PHM dashboard

- Staff members / board members / chairpersons of a PCZ or general practitioners association;
- Local government officials involved in healthcare;
- Healthcare professionals working in primary or secondary care withing the living lab;
- ICT professionals involved in dashboards at a local level;
- Coordinators of hospital networks;
- Population health managers from the living labs.

#### 3. Participants should possess the following prior knowledge:

- **Healthcare Systems**: Basic understanding of how healthcare systems operate, including healthcare delivery models, financing mechanisms, and regulatory frameworks.
- **Epidemiology**: Basic understanding of disease spread within populations, including risk factors and disease patterns.
- **Data Analysis and Management**: Basic knowledge of collecting, analysing, and interpreting health-related data to identify trends and disparities.
- **Public Health Strategies**: Familiarity with public health strategies aimed at promoting health, preventing disease, and improving accessibility.
- **Interventions**: Basic knowledge of population-level programs and their implementation.
- **Health Policy**: Basic insight into healthcare policy at local, national, and Flemish levels.
- **Interdisciplinary Collaboration**: Basic knowledge of collaborating with various stakeholders, such as healthcare providers, policymakers, social organizations, and local authorities, to address complex health challenges.
- **Health Promotion and Education**: Basic understanding of implementing health promotion strategies.

### Supplemental material 2 Topic guides

#### Focus group 1

**Participant Presentation:**

- Name and Surname
- Function
- Current Interaction with PHM: How do you currently engage with Population Health Management (PHM)?

**Situation of PHM in Daily Practice:**

- Explanation by the Moderator: An overview of what a dashboard is, including the distinction between aggregated data and linked individual data.
- Example Demonstration: Show an example, such as a diabetes dashboard used in a general practice setting.

**Vision Around PHM:**

- Expectations and Needs: What are your expectations and needs regarding PHM?
- Application in Daily Practice: How do you envision using this in your daily practice?
- Level of Need: At what level is PHM needed (e.g., individual, organizational, regional)?
- Preconditions for Feasibility: What preconditions are necessary for PHM to be feasible in practice?
- Bottlenecks and Enablers: What are the main challenges and facilitators for implementing PHM?

Steenkamer's Collaborative Adaptive Health Network (CAHN) Spiral. Parameters and

Functionalities: For each item in the CAHN spiral, discuss which parameters are needed and what functionalities the dashboard should have. How would you like these to be visualized?

- Population Identification:
- Quintuple Aim Measurement: Discuss the measurement of health parameters, quality of care, healthcare costs, provider welfare, and equity.
- Risk Stratification: How should risk stratification be addressed?
- Interventions Around Citizens: What interventions should be designed around the needs of citizens?
- Quintuple Aim Result Measurement: How should the results of the Quintuple Aim be measured?
- Process Improvement: What aspects of the process need improvement?
- Benchmarking Points of Interest: What are the key considerations for benchmarking?

**User Profile:**

- General User Profile Description: Provide a general description of the users.
- Population Manager Profile Description: Provide a specific profile description for population managers.

### Focus group 2

**Participant Presentation:**

- Name and Surname
- Function
- Current Interaction with PHM: How do you currently engage with Population Health Management (PHM)?

**Feedback from the First Focus Groups:**

- Summary of Insights: Present key feedback and insights gathered from the initial focus groups.

**Presentation of the Mock-up Dashboard Draft Slides:**

- Focus Areas:
  - General Exploration: Overview of general features and functionalities.
  - Risk Stratification and Selection of Interventions: How the dashboard addresses risk stratification and intervention selection.
  - Research Community: Features relevant to the research community.
- User Types:
  - Population Health Manager
  - Expert in Population Health Management

**Applicability of the Mock-up Dashboard:**

- Meeting Expectations: Does the mock-up dashboard meet your expectations?
- Usage: How would you use this dashboard in your practice?
- Barriers and Enablers: What are the potential barriers and enablers to its implementation?
- Preconditions: What preconditions are necessary for successful implementation (e.g., training, financing, networking, and connections with the micro level)?

### Supplemental material 3 Dashboard requirements as identified by the focus groups

#### DATA

**Availability of data (sources) – Types, sources, and levels of available data**

- **Raw data** (underlying dashboard data)
- **Level**: primary care zone, city / town, neighbourhood, district, local government level, zip code, practice
- **Types** (quantitative and qualitative)
  - **Person characteristics**: age, BMI
  - **Individuals' health data**: BMI, kidney function, vaccination status (influenza vaccine, pneumococcal vaccine, COVID vaccine), therapy adherence, indication for drug prescription, QALY's, infectious vector diseases
  - **Socioeconomic data**: increased compensation, social map, vulnerability, domestic help / nursing care, income, education, origin, independent living difficulty
  - **Environmental factors**: heat stress, air pollution
  - **Data related to health and welfare professionals**: well-being of care professionals (including workload, administrative burden), experiences (of neighbourhood workers), FTE by dashboard theme
  - **Population health management analyses**: costs and expenses, health outcomes in mortality, history of executed evidence-based interventions, process and outcome indicators (defined by expert panel), bottom-up data (data obtained following interventions conducted upon dashboard analysis)
- **Sources**: IMA, Sciensano, social service of hospital, public centre for social welfare, health insurance
- **Inventory of existing databases**

| Facilitators | Barriers |
|---|---|
| • Absolute and relative metrics<br>• Defining the denominator<br>• Data entry by non-dashboard-users (to reduce data sharing barrier)<br>• Governance for selecting new dashboard themes and data | • Data-unavailability leads to non-determinability of indicators<br>• Data-unavailability at low geographical level<br>• Barriers to data-sharing: GDPR, population reluctant to share data, individuals revoking data sharing consent<br>• Absence of contextual (soft) data |

**Quality of data (sources) – Correctness and completeness**

- Up-to-date data

| | |
|---|---|
| • Dense data collection | |
| Facilitators | Barriers |
| • Involve data providers to improve registration quality<br>• Require researchers to format data for immediate dashboard use | • Perceived data inaccuracies lead to user drop-off<br>• Limited or unclear coding in the electronic health record (or other data sources) restricts data inclusion in dashboard. |
| **Interoperability of data (sources) – Seamless data combination** | |
| • Integrate data into a federated data network from various sources (e.g. hospitals, GPs, pharmacists, paramedics)<br>• Standardize dashboard architecture via consistent data definition (over time and area), ensuring operability across regions (e.g. national architecture with regional interfaces for Flanders/Wallonia) | |
| Facilitators | Barriers |
| / | • Intraorganizational data collection and preservation<br>• Data from different sources covering various regions complicates demarcation<br>• Data protection regulations hinder linking data from different sources |
| **USERS** | |
| **Role – Dashboard user profiles and their role(s)** | |
| • **Practice level** (e.g. general practitioners, pharmacists, ...) - Profiles with population health management function on practice level should analyse data, visualize results, and determine focus areas.<br>• **Population health manager** (on neighbourhood level or meso level)<br> ○ Manage dashboard and send notifications when issues arise<br> ○ Engage with healthcare and welfare workers (= neighbourhood consultation, multidisciplinary team of general practitioners, pharmacists, nurses, social government agency, directors of schools in the neighbourhood, physiotherapists, ...) relevant to the notification<br> ▪ 1. Manager explains data findings<br> ▪ 2. Gather neighbourhood team experiences about the topic (= combining quantitative and qualitative data)<br> ▪ 3. Evaluate potential causes<br>• **Clinical support manager** - Create a strategic care plan using dashboard data |  |

- **Primary care zone** – Collaborate with health professionals and local government to intervene as needed
- **Data-analyst**
  - Conduct in-depth analysis (e.g. scientific research)
  - Supporting dashboard users in: 1. proving data-based standpoints (of healthcare providers); 2. impact evaluation (incl. QALY calculation); 3. identifying correlations; 4. Interpreting data.
- **Policy makers**
  - Monitor domain-specific data to guide policy decisions
  - Prioritize interventions based on parameter alarms
- **Government** - Dashboard management

| Facilitators | Barriers |
|---|---|
| • Stakeholder cooperation (combination of different 'roles' depending on the topic) to gain in-depth knowledge on certain themes and map data to their context | • Fear of government control/punishment based on data (benchmark fear)<br>• Insufficient staff to work with dashboard (e.g. lack of time) |

Dashboard accessibility – (Partial) data access

- Align data levels (e.g. primary care zone, neighbourhood) and types (e.g. QALY's, independent living difficulty) with roles
- Data-analysis supporting features (e.g. filters to place on top of data) only accessible to trained users

| Facilitators | Barriers |
|---|---|
| • Single sign-on from medical software | • Misinterpretation of data available in the open access dashboard (e.g. by journalists or other (untrained) dashboard users with insufficient knowledge) |

**Training in dashboard use – Supporting correct and smooth dashboard use**

- E-learning: covering usage (what to find where, and how) and interpretation based on examples and exercises

| Facilitators | Barriers |
|---|---|
| • Tailor training to dashboard user profiles | x |

**ANALYSIS**

**Population health analysis – Evaluation of population health by dashboard (user) for identification and prioritization of challenges**

- **Analysis provided by dashboard**
  - **Highlight**
    - **When:** deviating parameter
    - **Content:**
      - Key indicators and comparisons for chosen region (health, social, economic)
      - Most affected regions for chosen theme(s)
      - Blind spots
    - **Customization:** user can customize additional highlights based on goal, target group, and theme
- **Analysis performed by user**
  - **Theme <-> region / target group:** starting from a theme, look at regions, or starting from a region, look at themes
  - **Insight into profiles behind metrics**
  - **Identify of blind spots**
  - Possibility to **add filters to maps**
  - **Benchmarking** to other (international) cities/practices/neighbourhoods with similar demographic and/or social characteristics
    - Dashboard suggests region(s) with which to compare
  - **Trends in time**
    - Global evolution
    - For own population/region
  - **Impact evaluation:** Based on executed actions and simulations
  - **Evolution of pyramid**
    - Detect switches of strata in the population and investigate causes / context / details
    - Make predictions about the evolution of the population within the pyramid
  - **Care program analysis**
  - **Determination of optimal budget spending**
- **Environment for data-analysts**

| Facilitators | Barriers |
| --- | --- |

<table>
<tr><td>• Cooperation between different stakeholders (e.g. healthcare and welfare workers + (local) policy + researchers) to gain in depth knowledge about certain themes and the population as well as to map the context behind the data (composition of team depends on the topic)<br>• AI supports insights and visualizations<br>• ‘Info’: support correct data analysis/interpretation (e.g. time frame of data collection)<br>• Space for personal notes to consider contextual factors in data interpretation<br>• Graph legend<br>• Visual representation of main message<br>• Sliders<br>• Search function<br>• Modular: user can prioritize key information and save layout<br>• Simple, intuitive visualization with low cognitive load (no data-analyst needed)</td><td>• Missing (contextual) data leads to skewed analysis (e.g. diplomats with separate health systems are counted incorrectly)<br>• Contextual factors affecting intervention outcomes, leading to incorrect impact evaluation<br>• Difficult to determine causality<br>• Fear of benchmarking with exceptional practices<br>• Political cherry picking<br>• Patients who continue to visit their GP, but do visit their local pharmacist gives an incorrect picture of what the GP prescribes and what the pharmacist delivers<br>• Data linking on individual level is often impossible<br>• Searching websites for additional info<br>• Complex dashboard leads to incorrect interpretation<br>• Data not immediately available in format suitable for immediate use (e.g. many clicks needed)<br>• Risk of assessment / evolution toward funding instrument may trigger gaming<br>• Highlights alone lead to little change (need support and solutions)<br>• Unclear how to identify blind spots</td></tr>
<tr><td colspan="2">Stratification – Theme-based classification of individuals based on the level of their condition</td></tr>
<tr><td colspan="2">• Classify individuals, belonging to a level / patient population of a (healthcare) professional, by theme into strata based on:<br>    ○ Severity of (health) theme (risk stratification)<br>    ○ Social factor(s) (socioeconomic stratification), then perform risk stratification per socioeconomic stratum<br>• Number of strata is theme specific<br>• Pyramid representation</td></tr>
<tr><td>Facilitators</td><td>Barriers</td></tr>
</table>

| | |
|---|---|
| • AI can use scientifically defined algorithms to stratify profiles based on personal data, providing initial population stratification insights WITHOUT considering the context<br>• Experts set strata alignment criteria | / |

**Intervention selection – Action proposals for addressing data-based observed challenges**

- **Define 'intervention'**
- **Prioritize interventions based on alarms**
- **Who -** Suggestions by multidisciplinary expert panel (e.g. scientists, local policy) and dashboard users
- **How - Suggestion mode**
    - Respond to challenges identified through data
    - Address blind spots
    - Consider budget, system burden, number needed to treat and return on investment
    - Per stratum
- **What - Suggestions**
    - Implemented interventions with info on context and impact to evaluate whether the intervention makes sense in the new context
    - Combination of activities
    - General (not subdivided into 1st, 2nd and 3th line)
    - National and local initiatives
    - Link interventions to prevention workers for support

| Facilitators | Barriers |
|---|---|
| • Provide expert support to low scoring care centres instead of punishment<br>• Government should allocate a 2-3% budget margin for targeted efforts based on dashboard data, ensuring people feel empowered to analyse and act on the data.<br>• Indicate impact of proposed intervention(s) using past similar situations<br>• Link proposed interventions to scientific evidence | • Interventions may conflict with local policies<br>• Linking the wrong people for intervention support (e.g. people who have never been in practice) |

| EVALUATION |
| --- |
| 1. User-centred-evaluation (questionnaire) → Who have used and/or are still using the dashboard, and why?<br>2. Why did people drop out?<br>3. Feedback collection |
